# Hierarchical Clustering of Canonical Retinal–Biopsychosocial Covariation Patterns Reveals Distinct Psychosis Subgroups: A Data-Driven Study from the UK Biobank

**DOI:** 10.1101/2025.08.18.25333918

**Authors:** Cemal Demirlek, Brendan Stiltner, Erik Velez-Perez, Victor Zeng, Steve Silverstein, Babatunde Aideyan, Paulo Lizano

## Abstract

**Background:** Retinal alterations and related health/cognitive profiles are reported in psychosis spectrum disorders (PSD), but it is unknown whether multivariate retina–biopsychosocial covariations can identify biologically distinct subgroups.

**Methods:** Layer-resolved retinal thicknesses were obtained from macula-centered optical coherence tomography (OCT) scans using UK Biobank data from 476 adults (238 PSD, 238 age- and sex-matched controls). Biopsychosocial measures included sociodemographic/economic, cardiometabolic, ocular, and cognitive measures. Canonical correlation analysis (CCA) was employed to characterize patterns of covariation between retinal and biopsychosocial measures. Statistically reliable retina–biopsychosocial covariations were hierarchically clustered to derive subgroups, which were compared across various profiles using general linear models with false-discovery rate (FDR) correction.

**Results:** CCA revealed a multivariate retinal–biopsychosocial association pattern (r=0.468, 95% CI=0.410-0.527, p<.0001) linking lower retinal thickness, primarily outer-layers with greater socioeconomic deprivation, higher BMI, poorer visual acuity and lower cognitive performance. Hierarchical clustering of CCA projections based on this multivariate pattern identified 2 internally stable groups. Patients in cluster 1 (retina-preserved, 43.7% PSD, 81.9% controls) were cytoarchitecturally and systemically similar to controls, except for thicker nerve fiber layer (RNFL, Cohen’s *d*=0.32, p_FDR_=0.0093). Cluster 2 (retina-impaired) had significantly more participants with PSD (76% PSD) and they exhibited lower thicknesses across total macula (*d*=-0.80, *p*_FDR_=5.64×10^−12^), RNFL (*d*=-0.39, *p*_FDR_=0.0009), and photoreceptor layers (*d*=-0.42 to -1.06, *p*_FDR_<.0001), and were also characterized by lower socioeconomic status (*d=*-0.71 to -1.35, *p*_FDR_<.0001).

**Conclusions:** Integrating layer-resolved OCT with biopsychosocial measures can noninvasively stratify PSD into biologically meaningful subgroups, underscoring the translational potential of retinal microstructure for precision psychiatry beyond symptom-based classifications.

## INTRODUCTION

As an accessible ontogenetic outpouching of the brain, the retina offers a non-invasive interface with the central nervous system. Imaging techniques such as optical coherence tomography (OCT) enable high-resolution, in vivo visualization of neural, synaptic, and photoreceptor changes (Supplementary-Figure 1) (1–6). A growing number of OCT studies report that psychosis spectrum disorders (PSD) patients, including schizophrenia (SZ) or bipolar disorder (BD), have thinner retinas. Several meta-analyses provide compelling evidence of decreased thickness—most consistently within the ganglion cell/inner plexiform layer (GC-IPL), outer nuclear layer, and retinal pigment epithelium (RPE)—while retinal nerve fiber layer (RNFL) findings remain mixed (7–11). Furthermore, retinal measures may reflect neurobiological changes shaped by both genetic (6,12–14) and environmental factors (15,16), offering insight into shared and distinct neurodevelopmental and neurodegenerative processes across the psychosis spectrum (17–20).

**Figure 1.**
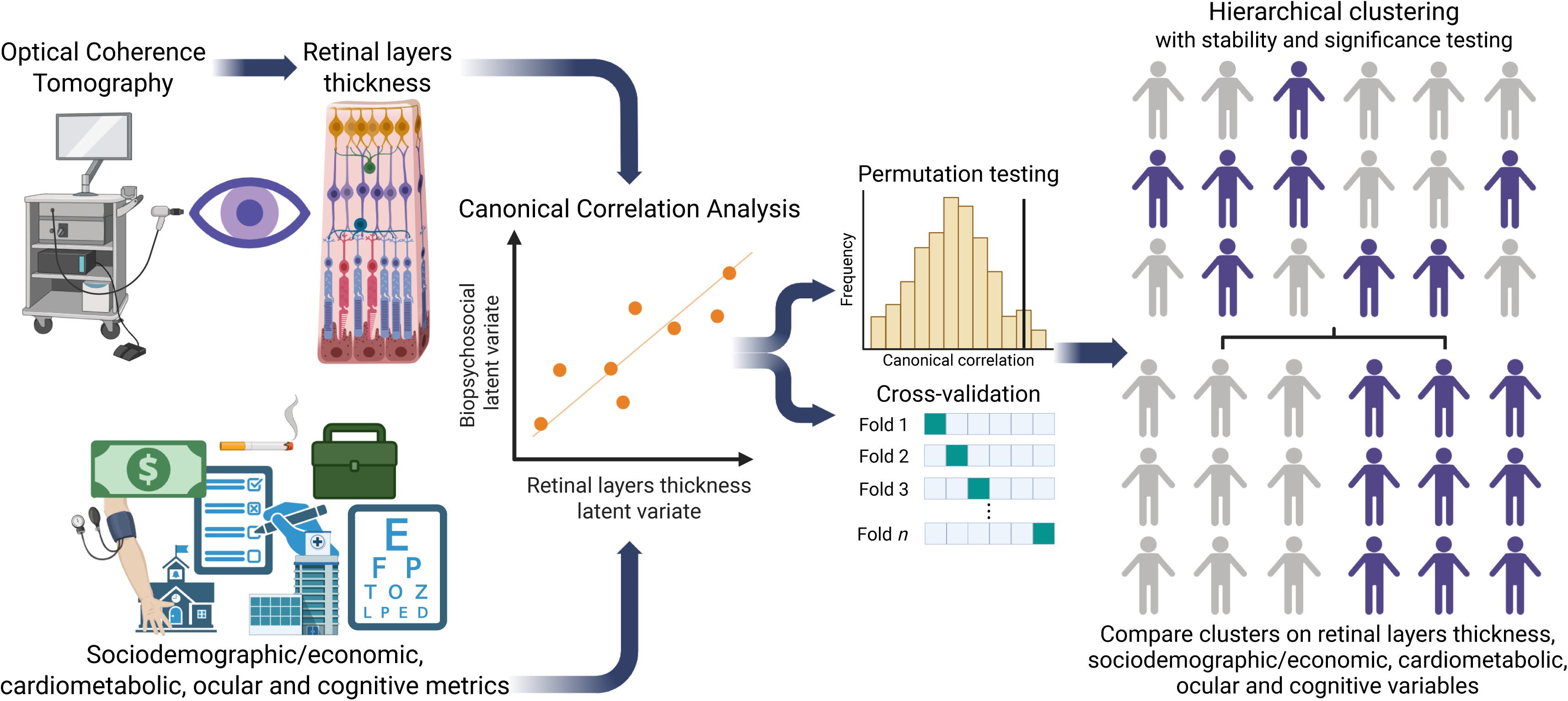
Study Design and Analysis Plan. Using data from the UK Biobank, optical coherence tomography was performed to image the thickness of retinal layers. Biopsychosocial variables, including sociodemographic/economic, health status, ocular metrics, and cognitive measures, were also assessed. Canonical correlation analysis was performed to identify modes of association between these two sets of variables. Then, using participants’ projections onto the latent variates, we hierarchically clustered these scores to identify clusters defined by retinal layer thickness and biopsychosocial metrics. General linear models were used to compare these clusters on these variables.

Retinal microstructure is shaped by a myriad of factors—including age, sex, race/ethnicity, material deprivation, systemic exposures (hypertension, obesity, smoking), visual acuity, and intraocular-pressure— many of which differ between patients and general population (21–25). Exploring how these factors interact can provide insight into complex relationships beyond the effects of the illness itself, comorbidities, environmental influences, and other health-related factors. However, most prior OCT studies do not consider these factors, relying on mean group differences, modest sample sizes, and rarely exploring the full retinal cytoarchitecture, including outer photoreceptor complex. Large, phenotypically rich, population-based resources (e.g. UK Biobank-[UKB]) therefore offer a platform for disentangling these interactions among retinal microstructure.

PSD, a construct that includes various psychotic disorders, shares a triad of positive symptoms, affective dysregulations, cognitive deficits, as well as excess mortality conferred through cardiometabolic comorbidity (26–28). Genome-wide association studies reveal extensive pleiotropy; neuroimaging shows overlapping gray-, white-matter, and connectivity abnormalities; neuropsychology shows overlapping cognitive dysfunctions between PSDs (29–32). Despite these commonalities, PSDs are also notoriously heterogeneous, blurring traditional diagnostic boundaries and motivating transdiagnostic approaches that seek common mechanisms while parsing homogeneous biologically defined subgroups within PSD (27,33–36). Initiatives such as the Bipolar-Schizophrenia Intermediate Phenotype (B-SNIP) have used cognitive and electrophysiology data to derive psychosis subgroups that cut across DSM categories, revealing clusters of latent patient groups with different neurobiology and prognosis (33,37). Retinal imaging—with its scalability, neural specificity, and biological plausibility (38,39)—has not yet been used for subgrouping. Therefore, beyond mean group differences, a critical question is whether retinal imaging can aid in stratifying the heterogeneity of psychosis and provide clues to distinct biological subtypes.

In this study, leveraging OCT and extensive phenotyping from the UKB, a datalZldriven workflow was adopted to (i) characterize multivariate associations between layerlZlresolved retinal thickness and sociodemographic, socioeconomic, health, ocular, and cognitive variables using canonical correlation analysis (CCA); and (ii) identify clusters by hierarchically clustering the resulting latent variate projections. We hypothesized that (a) a dominant CCA variate would couple lower thickness in specific retinal layers with poorer health indicators and cognitive impairments, and (b) this multimodal signature would delineate a PSDlZlpredominant subgroup exhibiting thinner retina relative to both other patients and healthy subjects (HC).

## METHODS AND MATERIALS

### Design, Participants, and Setting

Data from the UKB was utilized. The UKB comprises individuals aged 40–70 years, recruited across the UK between 2006-2010 (40). Participants with PSD—including SZ (ICD-10: F20–F29), BD (F30.2, F31), and major-depressive-disorder with psychosis (F32.3, F33.3) were included. Participants with eye/adnexal disorders (ICD-10; Chapter-VII), nervous system disorders (ICD-10; Chapter-VI), and other severe systemic illnesses were excluded. PSD participants were not excluded based on co-occurring mental/behavioral diagnoses. For HC, the absence of any diagnosed mental/behavioral disorders (ICD-10; Chapter-V) was required. Participants with duplicate entries, retinal scan from only one eye, missing information on age or birth-sex, or retinal thickness values considered outlier (Z-scores >+3 or <–3) were excluded. When available, data were matched across instances. After these criteria, 23,397 participants remained eligible. To reduce confounding effects of age and sex, frequency matching in a 1:1 case-control design was used, resulting in a final sample of 476 participants matched on age and sex. All participants provided written informed consent, and participants who later withdrew consent were excluded. The STROBE (41) and RECORD (42) guidelines were followed and the study was conducted in accordance with Declaration of Helsinki. The study was approved (No:11/NW/0382) by the Northwest Multicenter Research Ethics Committee.

### Retinal Imaging

Macula-centered retinal scans were obtained with Topcon 3D OCT 1000 Mk2 under mesopic conditions without pupil dilation. Both eyes were scanned using a protocol that captured 512 A-scans per B-scan across 128 horizontal B-scans arranged in 6x6mm raster pattern. Scans were segmented using Topcon Advanced Boundary Segmentation software (v1.6.1.1). Total macular retina and eight macular layers were extracted: RNFL, GC-IPL, inner nuclear layer (INL), inner nuclear layer to retinal pigment epithelium (INL–RPE), inner nuclear layer to external limiting membrane (INL–ELM), external limiting membrane to photoreceptor inner/outer segments (ELM–ISOS), inner/outer segments to RPE (ISOS–RPE), and RPE, following the segmentation protocols (43). Quality control included scans only with a quality score greater than 45 (UKB Data Fields 28553 and 28552) (44). Thicknesses of the central, inner, and outer subfields for the photoreceptor layers were extracted.

### Biopsychosocial Variables

Biopsychosocial variables spanning sociodemographic/economic, cardiometabolic, ocular, and cognitive domains were used. Sociodemographic/economic factors included age, sex, race/ethnicity, annual household income, and Townsend Deprivation Index (TDI)–indicator of material deprivation (45). General health and cardiometabolic indicators included body-mass-index (BMI), systolic blood pressure, tobacco-smoke exposure, and alcohol-use. For ocular health, best-corrected-visual-acuity (BCVA) in logMAR units, corneal hysteresis, and intraocular-pressure were used. Cognitive measures included processing speed via Reaction-Time test, fluid intelligence, and prospective memory via Shape test (46,47). Because of missing health, ocular, and cognitive measures (percent missing: range=0.21-12.4%, mean=4.3%, median=2.7%), analyses were conducted using random forest multiple imputation. The imputed results are reported here, while the supplemental results contain complete cases. Rubin’s rule was used for pooling estimates and calculating variance across imputations (48).

### CCA

To investigate the patterns of relationships between retinal layer thickness and biopsychosocial variables, CCA was performed (**Figure 1**). Thickness measurements included RNFL, GC-IPL, INL, and RPE, along with available subfield data (central, inner, outer) for INL-ELM, ELM-ISOS, and ISOS-RPE layers (6). Total macula and INL-RPE layers were excluded from the CCA because of their overlap with other layers. Biopsychosocial variables used were those available in UKB. First, variables were tested for collinearity and a stepwise approach to remove those with a variance inflation factor greater than 10, which removed the inner subfield of ISOS-RPE and INL-ELM and corneal-compensated intraocular-pressure. CCA projections for PSD and HC participants were compared in each imputation. CCA significance and reliability were quantified using permutation tests and cross-validation (49) (supplemental-methods).

### Hierarchical Clustering

Hierarchical clustering was conducted to identify subgroups. Using statistically significant and reliable CCA latent variate pairs, participants’ latent variate projections were hierarchically clustered by calculating Euclidean distances. The linkage method with the largest agglomerative coefficient was used for clustering. Hierarchical clustering was performed for each imputation—using CCA projections from that imputation—resulting in 10 cluster assignments. Participants were assigned to the cluster they were most frequently assigned to over the 10 imputations. If there was a tie, the silhouette score (50) was calculated for each imputation, then averaged across these imputations and assigned to the cluster with the highest average silhouette score. The average silhouette score across all participants was calculated to determine the ideal number of clusters. Cluster significance and stability were assessed following previously reported procedures (49). Significance testing constituted random sampling the covariance matrix of the latent variates’ projections (n=1000), calculating the silhouette score for each sample to create a null distribution, and comparing the silhouette score obtained with the data to this null distribution to compute a p-value. Cluster stability was assessed using the Jaccard similarity index (a value ranging from 0-1, where >0.75 indicates stable clusters) to compare the cluster assignments across bootstrapping (n=1000) (51). Lastly, median splits of the PSD sample were performed using total macula thickness, prospective memory, two latent variates’ projections, and the slope of the two latent variates. The effect size estimates were compared for these median splits to the clusters derived from hierarchical clustering, testing if our clustering procedure was better than median splits of retinal layer thickness and cognition (supplemental-methods).

### Cluster Differences

Because our goal was to dissect heterogeneity within PSD, not to subtype HC, all HCs were pooled into a single normative reference—despite a small retina-impaired minority—to retain statistical power. Clusters of PSD participants and HC were compared using general linear models, assessing differences in retinal layers thickness measures and biopsychosocial variables. Analyses were first conducted without covariates before using models adjusting for (i) age, (ii) race, and (iii) age and race. P-values were corrected for false-discovery rate (FDR) (52). All analyses were conducted between June 2024 and April 2025 using RStudio (v4.2), except for PSD-HC 1:1 (age & sex) matching (conducted using SPSS v29.0.2).

## RESULTS

### Participants

A total of 238 PSD participants were agelZl and sexlZlmatched to 238 HC. Both groups had identical mean and standard deviation for age (53.5±8.4 years, range=40–69) and sex distribution (47.1% male). Most participants identified as White (PSD=87.4%, HC=91.6%), see Supplementary-Table 1.

### CCA Results

The first CCA latent variate had a canonical correlation *r*=0.468 (95% confidence interval (CI)=0.410-0.527, **Figure 2A**). In each of the 10 imputations, participants with PSD had lower weighting onto the retinal layer thickness latent variate (Hedges’ *g* range: 0.64-0.66, *p*<9.23×10^−12^, *p*_FDR_<9.23×10^−12^) and the biopsychosocial latent variate (*g* range: 0.80-0.83, *p*<2.91×10^−17^, *p*_FDR_<2.91×10^−17^) than HC (**Figure 2B-C)**. The first latent variate pair was significant following permutation testing (*p*<0.0001, **Figure 2D**) and similar canonical correlations over cross-validation (training-*r*=0.475, testing-*r*=0.338; **Figure 2F**). This first retinal imaging latent variate was defined primarily by outer and central subfields of ISOS-RPE, ELM-ISOS, and INL-ELM layers (**Figure 2G**), while the first biopsychosocial latent variate was informed by household income, prospective memory, BMI, race, TDI, and visual acuity (**Figure 2H**, see supplement for unimputed data).

**Figure 2.**
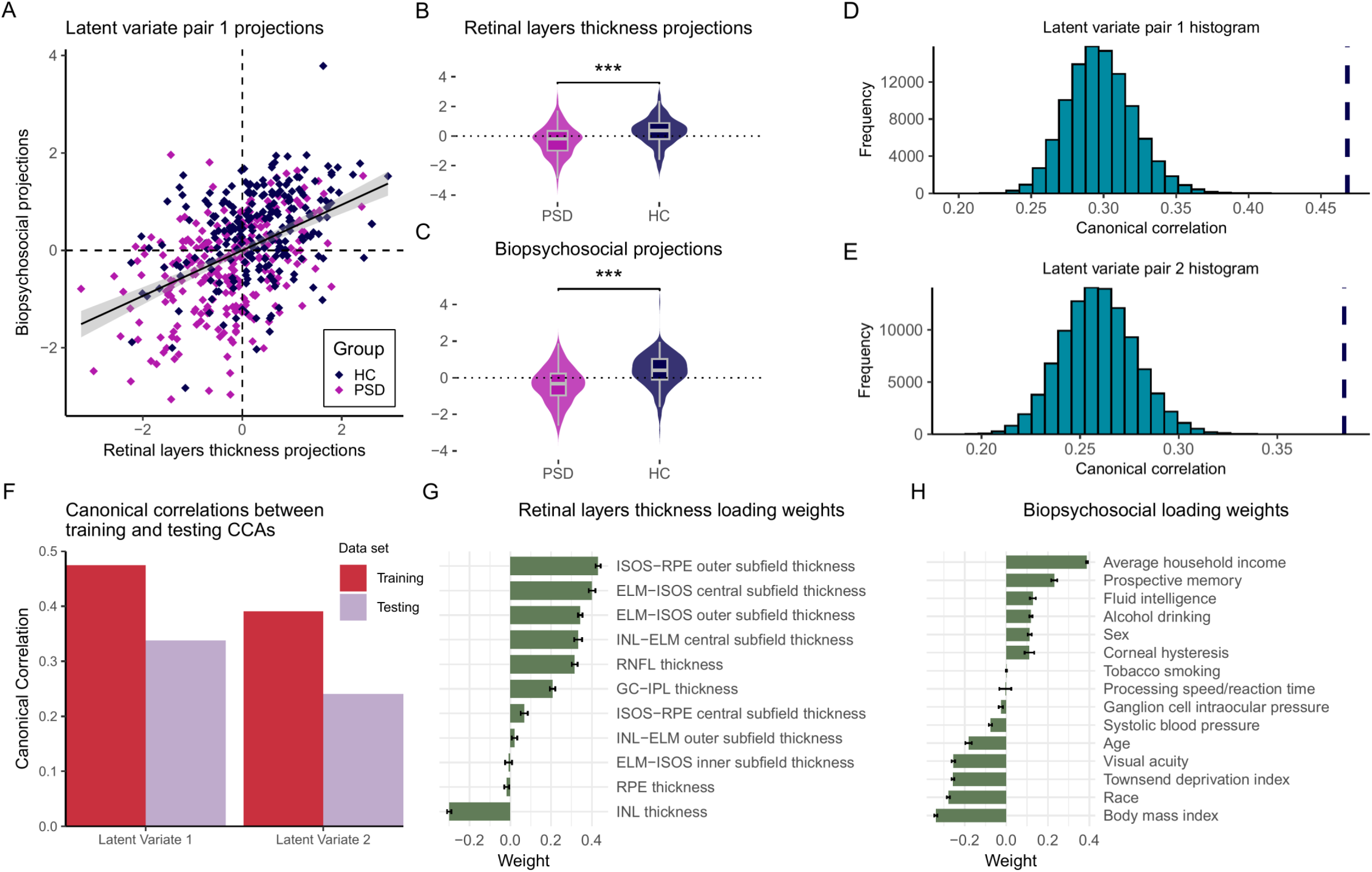
Canonical Correlation Results. **A.** The first latent variate identified by CCA (r=0.468). **B-C.** Participants with PSDs exhibited lower loadings onto the retinal layers thickness and biopsychosocial latent variates. **D-E.** Permutation testing found the first canonical correlation latent variate was significant (*p*<0.0001) as was the second latent variate (*p*<0.0001). While latent variate pairs 3-6 were also significant (*p*<0.01), they displayed smaller canonical correlations. Moreover, latent variate pairs 7-11 were not significant following permutation testing (*p*>0.05) (latent variates 1 and 2 are shown). **F.** Cross-validation showed latent variates 1 and 2 were reliable (latent variate 1 training−testing difference=0.137; latent variate 2 training−testing difference=0.15), with a drop for the remaining latent variates (latent variate 3 training−testing difference=0.239; latent variate 3 not shown). **G-H.** Average latent variate loading weights for the first latent variate for retinal imaging and biopsychosocial loading weights variables, respectively. 95% Confidence interval shown. **Note**. HC=Healthy controls; PSD=Psychosis spectrum disorders; ISOS=Inner/outer segments; RPE=Retinal pigment epithelium; ELM=External limiting membrane; INL=Inner nuclear layer; RNFL=Retinal nerve fiber layer; GC-IPL=Ganglion cell–inner plexiform layer.

### Hierarchical Clustering

Given the significance and reliability of the first latent variate pair, its projections were used for hierarchical clustering. Results indicated a 2-cluster solution best represented the distribution of the latent variate projections (**Figure 3A**). The average silhouette score was highest for 2 clusters (silhouette score=0.412, **Figure 3B**) and applying these cluster assignments to the plot of average CCA projections demonstrated 2 clusters (**Figure 3C**). The clustering was significant following significance testing (*p*=0.015, **Figure 3D**) and stable over bootstrapping (mean Jaccard indexes: Cluster 1=0.85, Cluster 2=0.79). Cluster 1 comprised 299 participants (n_PSD_=104, 43.70%; n_HC_=195, 81.93%) while cluster 2 contained the remaining 177 participants (n_PSD_=134, 56.30%; n_HC_=43, 18.07%). Cluster 2 had significantly more participants with PSD than HCs (χ^2^ =72.85, *p*<0.0001). The effect size estimates of those derived from hierarchical clustering compared to the median splits showed that the hierarchical clustering procedure produced estimates capturing similar or greater differences than the median splits (supplemental results).

**Figure 3.**
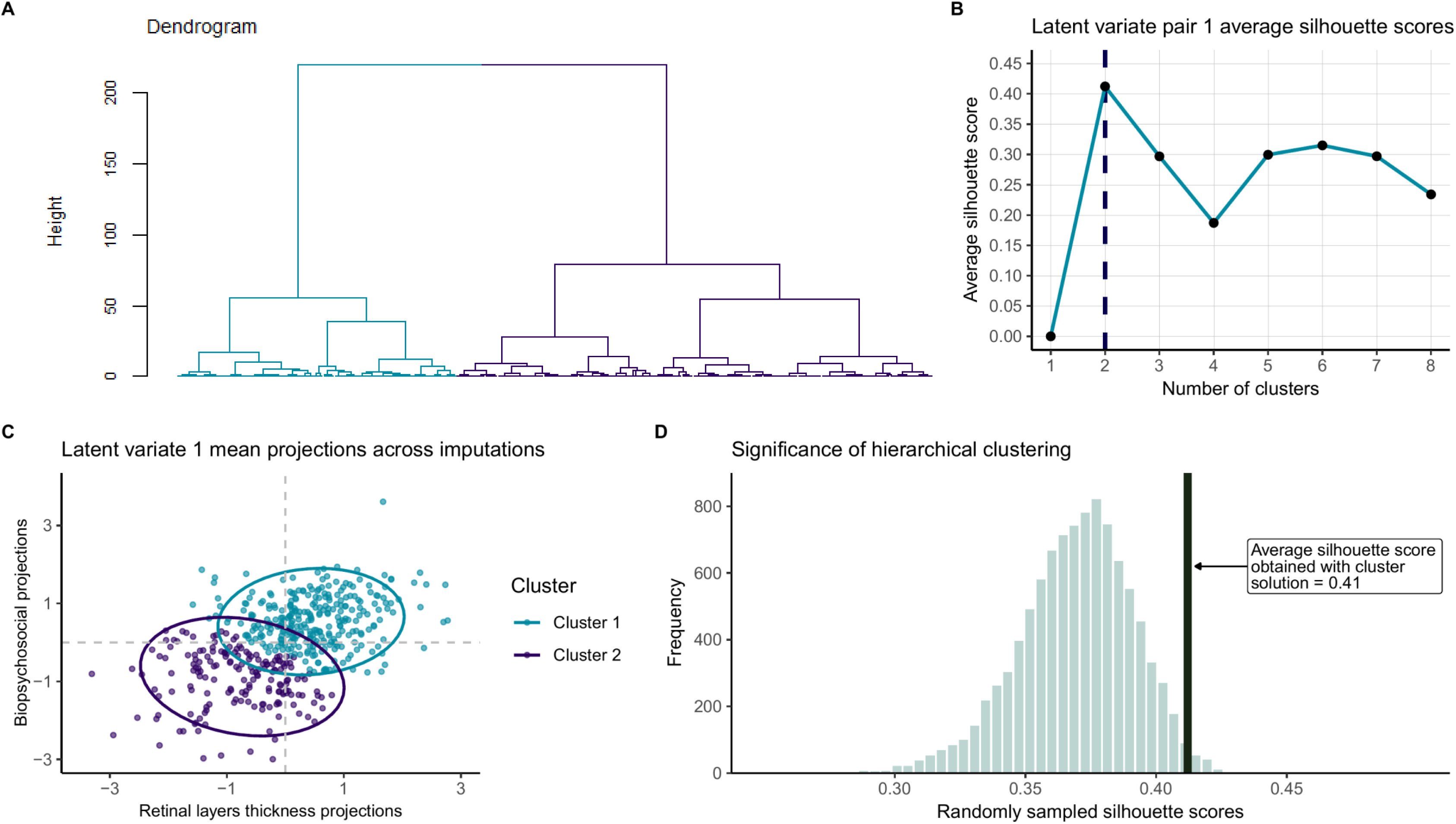
Hierarchical clustering results. **A.** Representative dendrogram of imputation 1, showing a representative example of the clustering of the first latent variate projections. Light blue branches correspond to cluster 1; purple branches correspond to cluster 2. **B**. Average silhouette scores calculated following an increasing number of clusters from the majority vote clustering. **C**. Plot of retinal layer thickness projections and biopsychosocial projections, with cluster assignments. Ellipses show 95% confidence interval for mean projections. **D**. Histogram showing frequency of simulated silhouette scores from random sampling and silhouette score observed from clustering solution.

### Differences in Retinal Layers Thickness Between Clusters

Compared to cluster 1, cluster 2 PSD exhibited thinner total macula, RNFL, INL-RPE, INL-ELM, ELM-ISOS, and ISOS-RPE (Cohen’s *d=*-0.50–-0.83, all *p*_FDR_<0.001), along with thicker RPE layer (*d*=0.35, *p*_FDR_=0.0078). Additionally, all photoreceptor subfields were thinner (*d*=-0.39–-0.95, all *p*_FDR_<0.006). GC-IPL and INL layers showed no differences (Figure 4 and supplement results.).

**Figure 4.**
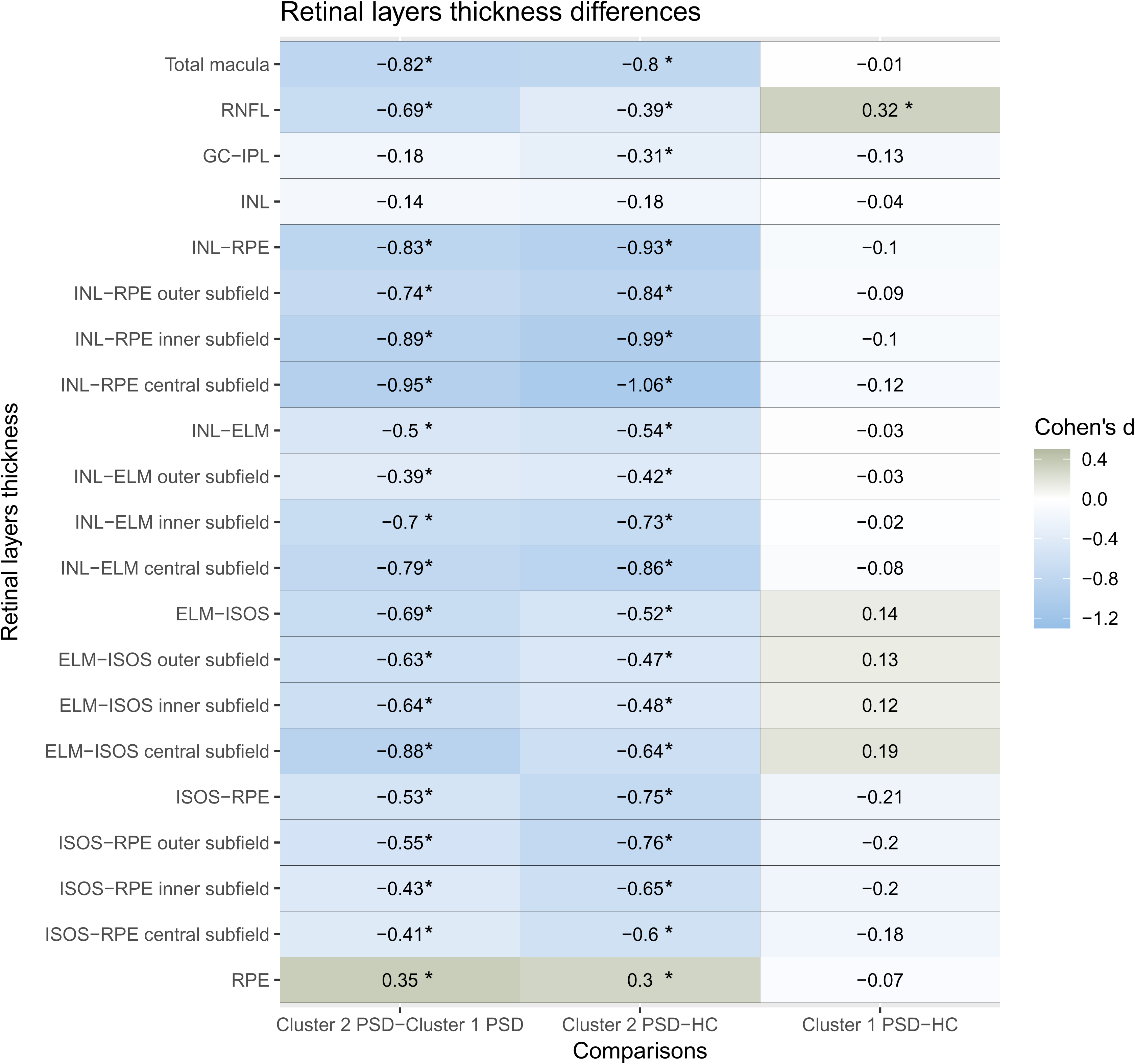
Differences in retinal layers thickness between clusters. Asterisks indicate comparisons significant following false-discovery rate correction, *p*_FDR_<0.05. **Note**. RNFL=Retinal nerve fiber layer; GC-IPL=Ganglion cell–inner plexiform; INL=Inner nuclear layer; RPE=Retinal pigment epithelium; ELM=External limiting membrane; ISOS=Inner/outer segments; PSD=Psychosis spectrum disorders; HC=Healthy controls.

Similarly, cluster 2 PSD compared to HC exhibited significantly thinner total macula, RNFL, GC-IPL, INL-RPE, INL-ELM, ELM-ISOS, and ISOS-RPE (*d*=-0.31–-0.93, all *p*_FDR_<0.008), along with thicker RPE (*d*=0.30, *p*_FDR_=0.0079). Additionally, all photoreceptor subfields were also thinner (*d*=-0.42–-1.06, all *p*_FDR_<0.0003), with INL showing no difference. Participants in cluster 1 PSD did not show differences from HC, except for thicker RNFL (*d*=0.32, *p*_FDR_=0.0093).

### Differences in Biopsychosocial Variables

Patients with PSD in cluster 2 differed from those in cluster 1 on age (*d*=0.45), race (*d*=0.50), income (*d*=−1.12), and TDI (*d*=0.71) (all *p*_FDR_<.005), but not sex, health indicators (BMI, blood pressure, alcohol use, tobacco smoking), ocular metrics (visual acuity, intraocular-pressure, corneal hysteresis), and cognitive functions (processing speed, reaction time, prospective memory). Although fluid intelligence (*d*=−0.75, *p*=0.045, *p*_FDR_=0.17) and prospective memory (*d*=−0.70, *p*=0.059, *p*_FDR_=0.19) were significant before FDR correction (Figure 5 and supplement results).

**Figure 5.**
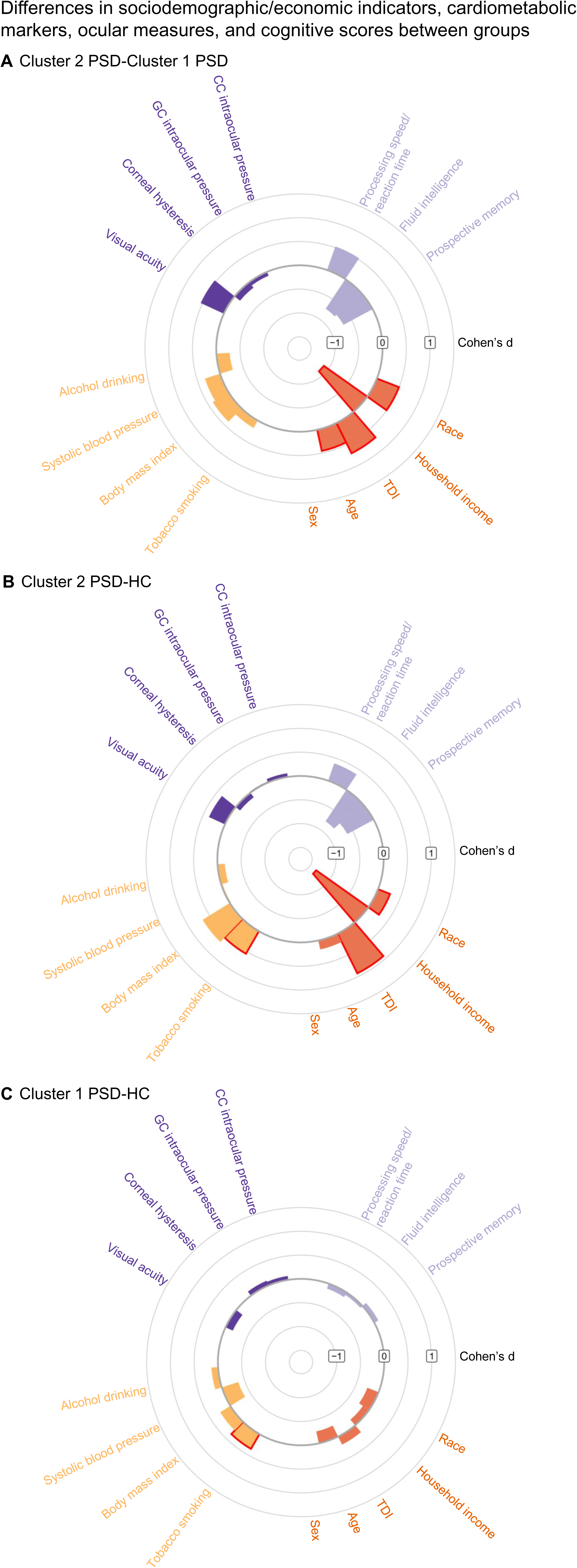
Differences in clinical metrics between clusters. Bars with red borders were significant following p_FDR_<0.05. A higher score on the Townsend Deprivation Index (TDI) indicates a higher level of material deprivation. Note. PSD=Psychosis Spectrum Disorders; HC=Healthy Controls; GC=Ganglion cells; CC=Corneal-compensated.

Patients in cluster 2 PSD differed from HC on race (*d*=0.27, *p*_FDR_=0.034), household income (*d*=−1.35, *p*_FDR_=1.11×10^−26^), TDI (*d*=0.93, *p*_FDR_= 2.41×10^−15^), and exposure to tobacco smoking (*d*=0.55, *p*_FDR_=1.42×10^−5^), but not age, sex, systolic blood pressure, alcohol consumption, or ocular measures (visual acuity, intraocular-pressure). Although fluid intelligence (*d*=−0.79, *p*=0.018, *p*_FDR_=0.088) and prospective memory (*d*=−0.63, *p*=0.058, *p*_FDR_=0.19) were significant before FDR correction.

Patients in cluster 1 PSD did not show significant differences from HC, except for exposure to tobacco smoking, where patients had higher rates of tobacco use (*d*=0.36, *p*_FDR_=0.045).

## DISCUSSION

In this data-driven study of retinal imaging in PSD, using CCA to link retinal microstructure with various biopsychosocial and health-related variables, we identified a dominant multivariate association pattern: thinner retinal layers —especially photoreceptors— covaried with indicators of material deprivation and poorer general health. Hierarchical clustering of participants based on this multivariate pattern delineated 2 internally stable clusters. PSD patients in cluster 1 (retina-preserved) were cytoarchitecturally and systemically similar to HC, except for thicker RNFL. Cluster 2 (retina-impaired) included most PSD patients, and these patients showed thinner retinal layers—except GC-IPL and INL— alongside material deprivation compared to both retinal- preserved PSD and HC groups.

Current findings both replicate and extend prior research examining associations between retinal microstructure and cognition in PSD. Boudriot et al (12), who used partial least squares analysis in SZ spectrum, identified a significant retinal-clinical-cognitive mode, mainly in inner layers. In contrast, our CCA identified a pattern involving both outer and inner layers and biopsychosocial variables, including socioeconomic and cognitive measures. Notably, outer retinal layers—particularly photoreceptors—contributed strongly to the canonical loadings in our sample, suggesting potentially distinct biological mechanisms not captured in prior work. By further applying hierarchical clustering to these canonical covariates, we delineated biologically meaningful subgroups within PSD, moving beyond conventional group-level, symptom-based analyses. The potential of retina to define biologically meaningful PSD subgroups aligns broadly with multimodal subgrouping efforts like B-SNIP. The retina-preserved group resembles B-SNIP’s ‘biotype-3’, with relatively intact neurostructural and cognitive profiles, whereas the retina-impaired group resembles ‘biotype-2’, marked by pronounced biological changes and intermediate cognitive deficits (54). ‘Biotype-1’ equivalent group may be absent here because of UKB’s comparatively healthier sample (53). Although not fully overlapping, retinal measures add complementary context to psychosis subtyping.

Among retinal layers, GC-IPL has consistently been reported as thinner in previous OCT studies of PSD. In our multivariate model, GClZlIPL contributed modestly to cluster segregation compared to outer layers, and it was not significantly different between cluster 1 and 2 patients, suggesting that GC-IPL changes in psychosis may be more globally distributed across the spectrum, whereas outer retinal changes demarcate more specific biological subtype. Additionally, thinner GClZlIPL was reported in unaffected relatives (54,55) and was associated with SZ polygenic risk (56), supporting its candidacy as an endophenotypic marker. Although thinner GC-IPL was replicated across SZ and BD meta-analyses, RNFL findings were inconsistent (7–10). In the current study, increased RNFL thickness in retina-preserved cluster underscores PSD heterogeneity and helps reconcile prior inconsistent results. There are three potential hypotheses for the thicker RNFL observed in the retina-preserved group: 1) lower cumulative exposure tied to less deprivation (e.g., smoking, poor nutrition, metabolic disease), 2) premorbid neural reserve that confers resilience, or 3) chronic low-grade inflammation resulting in RNFL expansion. Disentangling these hypotheses requires a more integrated evaluation that includes blood-, retina-, brain-, and expo some-based assessments.

Moreover, a distinct PSD subgroup with pronounced decreased outer retinal thickness was more likely to experience material disadvantage and poorer health. These outer layers include dendrites of bipolar cells, inner and outer segments of photoreceptors, all of which are regions of high metabolic activity (57). Therefore, these layers are markedly sensitive to bioenergetic/mitochondrial stress, microvascular compromise, and inflammation^58^. The preferentially decreased thickness of these layers suggests potential vulnerability to systemic or environmental stressors, especially in socioeconomically disadvantaged individuals. Another UKB study identified genetic loci associated with thinner photoreceptor layers and demonstrated their links with increased risks for systemic cardio-pulmonary-metabolic conditions, as well as incident mortality (59).

Additionally, photoreceptor dysfunction in early psychosis was associated with childhood trauma and insulin resistance (15). These highlight the value of considering both biological and social factors in characterizing disease expression in PSD.

Importantly, retinal changes are known to precede clinical decline in neurological diseases (60–62). Extending this concept, measuring retinal thickness in PSD, or even those in clinical-high-risk and first-episode psychosis (63,64), may help identify those on a trajectory of greater biological vulnerability to certain factors (15). A retina-impaired signature could facilitate early risk stratification and provide an objective biomarker for tracking neuroprogression within the framework of precision psychiatry.

Although cognitive differences didn’t survive FDR correction, retina-impaired patients demonstrated larger effect-size deviations compared to both HC and retina-preserved patients, supporting the translational potential of retina as a low-burden biomarker for functional brain health. The retina-impaired cluster might represent a group where cognitive impairment is moderate, but where environmental factors have contributed to neuroprogression. Studies in clinical-high-risk and first-episode psychosis have shown no thinning in outer layers, while cognitive dysfunctions correlated with macular and GC-IPL thickness, but not with outer layers (60,61). In this context, cognitive deficits may reflect both neurodevelopmental and neurodegenerative vulnerabilities (62–64), while outer layer alterations may accumulate with disease progression. Ultimately, longitudinal retinal-biopsychosocial assessments are crucial to further clarify these relationships.

## Limitations

The cross-sectional nature and lack of data on medication use, illness duration, and clinical outcomes are key limitations. Future research incorporating longitudinal and multimodal data will provide deeper insights into the role of retina in PSD. Additionally, data-driven clustering can be sensitive to the methods, data structure, and sample (65), and further investigation is needed to refine optimal subtypes by incorporating different biomarkers, such as brain imaging, inflammation, or genetic profiles. Finally, this study’s generalizability is also limited due to UKB’s participant pool, which is predominantly older and less ethnically diverse.

## Conclusion

We identified multimodal retinal association pattern: thinner retinal layers—especially outer layers—were coupled with material deprivation and poorer general health. Based on this multivariate pattern, 2 biologically distinct data-driven subgroups of PSD were identified: one marked by thinner retina—especially photoreceptors—and socioeconomic disadvantage, and another defined by preserved retinal structure and cognitive performance, except for thicker RNFL. This work extends prior case–control OCT studies by demonstrating that retinal cytoarchitecture can uncover latent subgroups within the PSD population, paving the way for more precise stratification and personalized treatment strategies.

## Data Availability

All data are available at UK Biobank

## Acknowledgments

This research has been conducted using the UKB Resource under application number 75692. We gratefully acknowledge the UKB for providing open access to this invaluable resource, which supports student researchers and advances health research. Furthermore, this work uses data provided by patients and collected by the NHS as part of their care and support. We sincerely thank the participants who generously contributed their health data, as the UKB relies on the dedication of volunteers committed to fostering health research and promoting a healthier society. Lastly, we recognize the Biomedical Sciences Career Program at Harvard Medical School for facilitating collaboration among the authors.

