## Supplement for "Hierarchical Clustering of Canonical Retinal–Biopsychosocial Covariation Patterns Reveals Distinct Psychosis Subgroups: A Data-Driven Study from the UK Biobank"

**Supplementary Figure 1.** Illustration of retinal imaging with optical coherence tomography, showing the retinal layers and their components.

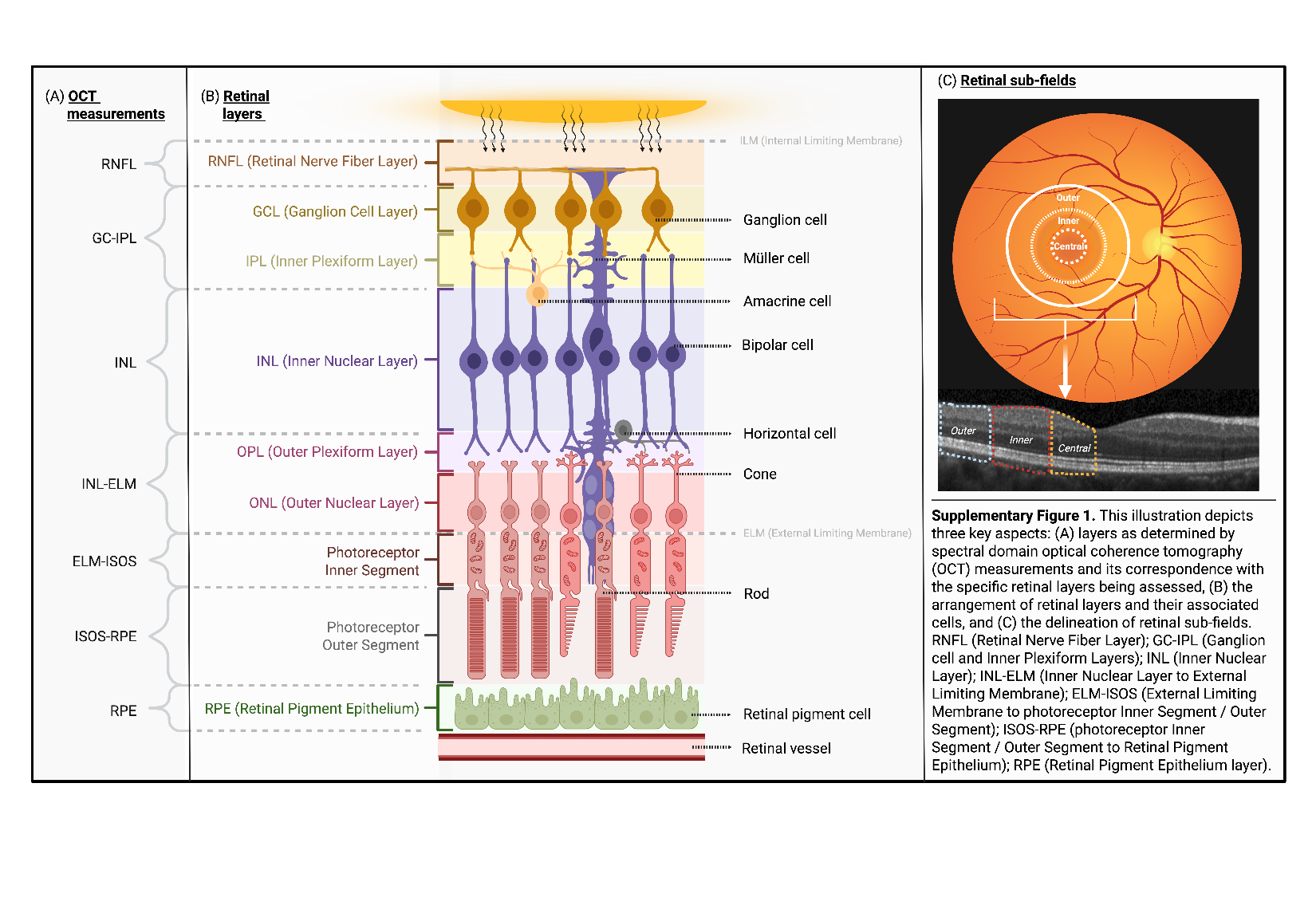

**Supplemental Methods**

*Imputation*

No retinal layer thickness or demographic data was missing. Systolic blood pressure (percent missing=0.21%), visual acuity (percent missing=0.21%), alcohol drinking (percent missing=0.63%), Body mass index (percent missing=0.84%), intraocular-pressure (percent missing=2.73%), processing speed/reaction time (percent missing=3.36%), fluid intelligence (percent missing=7.56%), corneal hysteresis (percent missing=11.76%), and prospective memory (percent missing=12.39%) were missing data. Data on tobacco smoking was also not missing. We chose to multiply impute data using the *mice* package in R with the random forest method with 10 imputations. Analyzes were performed on the 10 datasets and averaged.

*Canonical Correlation Analysis*

Because we had 10 imputed datasets, we scaled each imputed dataset, performed canonical correlation analysis (CCA), and saved the latent variate loading weights and projections from each imputation. To estimate within-imputation variance for averaging with Rubin’s rule, we performed bootstrap resampling within each imputation, stored the canonical correlation from the resampling, and calculated the variance. Then, the average canonical correlation was taken from the 10 imputations and the total variance was calculated following Rubin’s rule to obtain the standard error and 95% confidence intervals (CI). We compared the latent variate projections for the retinal layers thickness and biopsychosocial variates between participants with PSDs and HCs in each imputation. We performed Welch’s t-tests because variances between groups were not equal, but the projections were normally distributed and reported the range of results from the 10 imputations.

Next, we performed permutation testing by shuffling the rows of retinal layers thickness, creating simulated data. The data was simulated in the sense that the retinal layer thickness data no longer corresponded to the biopsychosocial data. This was done using the *rsample* package in R. We performed this shuffling of data 10,000 times, calculating the canonical correlation from each shuffled dataset and creating a null distribution. We calculated a p-value defined as the number of times the canonical correlation from the permutations were greater than the canonical correlation identified from averaging the canonical correlation across the 10 imputations. We performed this procedure in each of the 10 imputed datasets resulting in 10 p-values, which we averaged. When comparing our unimputed data to our imputed data, we performed this permutation test one time in the complete cases to calculate the p-value, comparing the number of times the 10,000 permutations canonical correlations were greater than the canonical correlation obtained from the real data.

Then, we conducted a cross-validation procedure. We used 10 folds, created with the *caret* package in R. This resulted in 90% of the sample’s data used for the CCA (training set), whose loading weights were applied to the data in the remaining 10% of the sample (testing set). The canonical correlation obtained in the training set and the testing set (by correlating the projections obtained by matrix multiplying the weights form the training set with the data from the testing set and taking the correlation of the retinal layer thickness latent variate and the biopsychosocial latent variate) were saved. This was repeated 100 times, creating different folds of participants each time. Then, this was performed in each of the 10 imputed datasets. Across all training and testing sets, from each fold, repetition, and imputation, the mean canonical correlations were taken for each latent variate. The more similar the canonical correlation is between training and testing sets, the more generalizable the latent variates are. Again, the same procedure was applied to the unimputed data but without performing it in multiple, imputed datasets.

*Hierarchical clustering*

Because we had 10 imputed datasets, we performed hierarchical clustering on the CCA projections to the latent variate pairs that were significant and reliable following our CCA testing in each imputation. We calculated the distance matrix for each latent variate pair using the Euclidean distance and testing average, single, complete, and Ward’s linkage methods. The linkage method with the highest agglomerative coefficient (ac) was used for clustering in each imputation. We combined the cluster assignments for each individual in the 10 imputations into a matrix and calculated each individual participant’s silhouette score in each imputation using the corresponding distance matrix. We tested k-numbers of clusters from k=2 through k=5. We averaged the silhouette scores for each participant to obtain a mean silhouette score for 2-5 clusters. The highest average silhouette score was used to determine what number of clusters to use moving forward. To obtain the final cluster assignments for those k-clusters, we returned to the matrix of cluster assignments from each imputation. We assigned participants to the cluster they were assigned to most frequently in the 10 imputations. For example, if k=3 and a participant was assigned to cluster 3 in 8/10 imputations, their final cluster assignment was cluster 3. If a participant tied for the most-often assigned cluster across the 10 imputations, we took the participant’s silhouette score from each imputation where they were assigned to the tied cluster assignment. We averaged the silhouette scores for the tied clusters and assigned them to the cluster that they had the higher, average silhouette score for. We refer to this approach as majority-vote clustering. It is based on the assumption that hierarchically clustering the CCA projections across imputations will produce clusters reflecting an underlying relationship between the retinal layers thickness and the demographic, ocular, health, and cognition that is the same across imputations.

We did not consider using consensus or ensemble clustering methods because they do not permit selecting a certain number of clusters to evaluate. Rather, they use a data-driven approach to ensemble multiple cluster assignments into one cluster assignment and the final k clusters is based upon the data (i.e., the clusters). Therefore, the final k clusters can be small (e.g., 2 or 3) or high (e.g., 7 or greater). We prioritized the interpretability of 2- to 5-clusters which we can set and determine via our majority-vote clustering approach.

For the resampling significance test and the bootstrapping stability test, we performed the approach described in the main test in each of the 10 imputations using the corresponding CCA projections. For the significance test, we compared the average silhouette score obtained from the final cluster assignment to the null distribution of silhouette scores obtained from hierarchical clustering the 10 imputation’s CCA projections using the resampling procedure. For the stability test, we used bootstrapping to resample CCA projections, perform the hierarchical clustering on each bootstrapped resample, and then compared the original cluster assignments to the bootstrapped cluster assignments with the Jaccard index.

In order to test if the clusters derived from hierarchically clustering the CCA projections merely reflect a split at the median of the retinal layers thickness-biopsychosocial dimension, we sectioned participants with PSDs into clusters by median-splitting the sample 5 different ways: using the median of the total macula thickness, the median of the prospective memory variable (chosen as a representative marker of the biopsychosocial variables, as it was one of the variables with the largest weight onto the biopsychosocial latent variate), the median of the retinal layers thickness latent variate projections, the median of the biopsychosocial latent variate projections, and the median of the retinal layer thickness-biopsychosocial dimension. To perform the median split of the retinal layer thickness-biopsychosocial dimension, we projected participants’ projections onto the slope of the best fit line of the latent variate pair’s projections. We performed general linear models comparing these median split clusters on different variables. For the total macula thickness median split clusters, we compared them to the prospective memory variable and compared the prospective memory median split clusters to the total macula thickness. For the retinal layers thickness latent variate projections median split clusters, we compared them to the biopsychosocial latent variate projections and the biopsychosocial latent variate projections median split clusters to the retinal layers thickness latent variate projections. For the median split clusters along the retinal layers thickness-biopsychosocial dimension, we compared them to projections to both the retinal layers thickness and biopsychosocial latent variates. These effect size estimates were compared to the effect sizes obtained using the clusters identified by hierarchically clustering the latent variate projections. We used Rubin’s rule for pooling cohen’s d across imputations.

For the unimputed, complete cases data, we performed the same analyzes but only once. The same procedure was performed: linkage methods were tested via their ac, the method with the highest ac was used for clustering, the highest, average silhouette score was obtained, cluster assignments for that number of clusters were obtained, and significance and stability testing were performed using those cluster assignments.

**Supplemental Results**

**Supplementary Table 1.** Sample characteristics.

| **Domains** | **Variables** | **HS (*n* = 238)**  *n* (%) | **PSD (*n* = 238)**  *n* (%) | ***p*** |
| --- | --- | --- | --- | --- |
| Socio-  demographic and economic  variables | Age | | | - |
|  | M ± SD | 53.5 ± 8.4 | 53.5 ± 8.4 |  |
|  | Range | 40, 69 | 40, 69 |  |
|  | Sex assigned at birth | | | - |
|  | Male | 112 (47.1) | 112 (47.1) |  |
|  | Female | 126 (52.9) | 126 (52.9) |  |
|  | Race / ethnicity |  |  | .086 ^b^ |
|  | White | 218 (91.6) | 208 (87.4) |  |
|  | Black | 2 (0.8) | 12 (5.0) |  |
|  | Asian | 8 (3.4) | 6 (2.5) |  |
|  | Multi-race / Others | 7 (3.0) | 7 (3.0) |  |
|  | Unknown | 3 (1.2) | 5 (2.1) |  |
|  | Townsend deprivation index | | | **< .001** ^c^ |
|  | M ± SD | -1.30 ± 2.95 | 0.59 ± 3.51 |  |
|  | Range | -6.26, 7.16 | -6.18, 8.34 |  |
|  | Annual household income | | | **< .001** ^b^ |
|  | Very low (< £18,000) | 41 (16.7) | 111 (46.6) |  |
|  | Low (£18,000 - £30,999) | 42 (17.1) | 36 (15.1) |  |
|  | Middle (£31,000 - £51,999) | 76 (30.9) | 56 (23.5) |  |
|  | High (£52,000 - £100,000) | 63 (25.6) | 21 (8.8) |  |
|  | Very high (>£100,000) | 24 (9.7) | 9 (3.8) |  |
|  | Unknown | 1 (0.4) | 5 (2.1) |  |
| Health  status  indicating  variables | PSD diagnoses | | | - |
|  | Schizophrenia | - | 105 (44.1) |  |
|  | Bipolar disorder | - | 97 (40.8) |  |
|  | Bipolar disorder with psychosis | - | 25 (10.5) |  |
|  | Major depressive disorder with psychosis | - | 11 (4.6) |  |
|  | Clinical | | | |
|  | Body mass index | 26.3 ± 4.1 | 28.6 ± 5.4 | **< .001** ^c^ |
|  | Systolic blood pressure (mmHg) | 136 ± 17 | 133 ± 17 | .064 ^c^ |
|  | Presence of cardio-metabolic disorder | 0 | 174 (73.1) | - |
|  | Cognition | | | |
|  | Processing speed (*Reaction time test*) ^a^ | 554.5 ± 126.8 | 577.1 ± 122.5 | .053 ^c^ |
|  | Fluid intelligence (*Fluid intelligence test*) ^a^ | 6.4 ± 2.2 | 5.4 ± 2.3 | **< .001** ^c^ |
|  | Prospective memory (*Prospective memory test*) ^a^ | 169 (81.3) | 140 (67.0) | **< .001** ^b^ |
|  | Behavioral | | | |
|  | Exposure to tobacco smoking (Yes) | 16 (6.7) | 51 (21.4) | **< .001** ^b^ |
|  | Exposure to drinking alcohol (Yes) | 227 (95.4) | 222 (94.5) | .652 ^b^ |
|  | Ocular | | | |
|  | Best corrected visual acuity | - 0.01 ± 0.16 | 0.01 ± 0.17 | .124 ^c^ |
|  | Corneal hysteresis | 11.0 ± 3.2 | 10.8 ± 1.7 | .436 ^c^ |
|  | IOP-cc (mmHg) | 31.5 ± 6.7 | 32.0 ± 6.9 | .353 ^c^ |
|  | IOP-gc (mmHg) | 31.5 ± 7.6 | 32.0 ± 7.1 | .475 ^c^ |
|  | Retinal image quality (QC) | 67.6 ± 9.6 | 67.8 ± 9.5 | .800 ^c^ |

Bold text: Indicates significant measures. PSD, psychosis-spectrum disorder. HS, healthy subjects. IOP-cc, corneal-compensated intraocular-pressure. IOP-gc, ganglion cell intraocular-pressure. QC, quality control.

^a^ Reflects a smaller sample due to incomplete task participation or missing values (Processing speed, n=7 in HS, n=9 in PSD; Fluid intelligence, n=13 in HS, n=23 in PSD; Prospective memory, n=30 in HS, n=29 in PSD).

^b^ Chi-square test or Fisher’s exact test

***Comparisons of median split clusters to clusters identified from hierarchical clustering***

When splitting participants with PSDs at the median of the total macula thickness, the two clusters did not differ with regard to prospective memory (d=0.26, *p*=0.47, *p*_FRR_=0.52). The hierarchical clusters did not significantly differ either with regard to prospective memory (d=−0.70, p=0.059, p_FDR_=0.084), though the effect was larger. Then, when splitting at the median of prospective memory, there was no difference in total macula thickness (d=0.20, p=0.60, p_FDR_=0.60), but the hierarchical clusters did differ by total macula thickness (d=−0.82, p=2.30×10^−9^, p_FDR_=2.30×10^−8^). When splitting the participants with PSDs at the median of the retinal layers thickness latent variate projections, they did not differ with respect to the biopsychosocial latent variate projections (d=0.66, *p*=0.070, *p*_FDR_=0.088) but they did with respect to the hierarchical clusters (d=−2.04, *p*=3.91×10^−7^, *p*_FDR_=1.30×10^−6^). Similarly, comparing the retinal layers thickness latent variate projections between the clusters split at the median of the biopsychosocial latent variate projections did not show a significant difference (d=0.72, p=0.051, p_FDR_=0.084) but the effect was significant in the hierarchical clusters (d=−1.62, p=2.87×10^−5^, p_FDR_=7.18×10^−5^). Lastly, the median split of the retinal layers thickness-biopsychosocial dimension produced significant differences in the retinal layers thickness latent variate projections (d=1.37, p=3.40×10^−4^, p_FDR_=6.80×10^−4^) and the biopsychosocial latent variate projections (d=2.29, p=2.60×10^−8^, p_FDR_=1.30×10^−7^), wherein the effect for the biopsychosocial latent variate was larger than those from the hierarchical clusters but the retinal layer thickness latent variate effect was smaller. To conclude, the clusters identified by hierarchical clustering revealed more pronounced or similar differences in retinal layers thickness and biopsychosocial variables than did those derived from different median splits of the data.

***Differences in retinal layers thickness between clusters***

When comparing participants with PSDs in cluster 2 to cluster 1, those in cluster 2 had significantly thinner total macula (*d*=−0.82, *p*=2.30×10^−9^, *p*_FDR_=9.67×10^−9^), RNFL (*d*=−0.69, *p*=1.10×10^−7^, *p*_FDR_=3.44×10^−7^), INL-RPE (*d*=−0.83, *p*=6.40×10^−10^, *p*_FDR_=3.1×10^−9^), INL-ELM (*d*=−0.50, *p*=0.00011, *p*_FDR_=0.00023), ELM-ISOS (*d*=−0.69, *p*=7.24×10^−7^, *p*_FDR_=2.07×10^−6^), and ISOS-RPE (*d*=−0.53, *p*= 5.18×10^−5^, *p*_FDR_=0.00011) layers and significantly thicker RPE layer (*d*=0.35, *p*=0.0047, *p*_FDR_=0.0078). When examining subfields of photoreceptor layers, cluster 2 also had thinner INL-RPE outer (*d*=−0.74, *p*=2.34×10^−8^, *p*_FDR_=8.18×10^−8^), inner (*d*=−0.89, *p*=3.29×10^−11^, *p*_FDR_=2.07×10^−10^), and central (*d*=−0.95, *p*=1.23×10^−12^, *p*_FDR_=1.10×10^−11^) subfields, thinner INL-ELM outer (*d*=−0.39, *p*=0.0030, *p*_FDR_=0.0053), inner (*d*=−0.70, *p*=7.85×10^−8^, *p*_FDR_=2.60×10^−7^), and central (*d*=−0.79, *p*=2.78×10^−9^, *p*_FDR_=1.09×10^−8^) subfields, thinner ELM-ISOS outer (*d*=−0.63, *p*=5.67×10^−6^, *p*_FDR_=1.37×10^−5^), inner (*d*=−0.64, *p*=5.51×10^−6^, *p*_FDR_=1.37×10^−5^), and central (*d*=−0.88, *p*=1.81×10^−10^, *p*_FDR_=9.50×10^−10^) subfields, and thinner ISOS-RPE outer (*d*=−0.55, *p*=2.73×10^−5^, *p*_FDR_=5.93×10^−5^), inner (*d*=−0.43, *p*=0.0011, *p*_FDR_=0.0020), and central (*d*=−0.41, *p*=0.0024, *p*_FDR_=0.0042) subfields layer thicknesses compared to cluster 1. The GC-IPL (*d*=−0.18, *p*=0.19, *p*_FDR_=0.25) and INL (*d*=−0.14, *p*=0.30, *p*_FDR_=0.37) layers were not different between cluster 2 and cluster 1.

Similarly, participants with PSDs in cluster 2 had significantly thinner total macula (*d*=−0.80, *p*=5.37×10^−13^, *p*_FDR_=5.64×10^−12^), RNFL (*d*=−0.39, *p*=0.00048, *p*_FDR_=0.00092), GC-IPL (*d*=−0.31, *p*=0.0047, *p*_FDR_=0.0078), INL-RPE (*d*=−0.93, *p*=2.22×10^−16^, *p*_FDR_=4.66×10^−15^), INL-ELM, (*d*=−0.54, *p*=9.41×10^−7^, *p*_FDR_=2.58×10^−6^), and ELM-ISOS (*d*=−0.52, *p*=2.83×10^−6^, *p*_FDR_=7.44×10^−6^), ISOS-RPE (*d*=−0.75, *p*=2.09×10^−11^, *p*_FDR_=1.47×10^−10^) and thicker RPE layers (*d*=0.30, *p*=0.0049, *p*_FDR_=0.0079) compared to HCs, while the INL layer was not different (*d*=−0.18, *p*=0.10, *p*_FDR_=0.15). When examining subfields of photoreceptor layers, cluster 2 also had thinner layer thicknesses for the INL-RPE outer (*d*=−0.84, *p*=8.30×10^−14^, *p*_FDR_=1.05×10^−12^), inner (*d*=−0.99, *p*=1.62×10^-18^, *p*_FDR_=5.11×10^-17^), and central (*d*=−1.06, *p*=3.40×10^-21^, *p*_FDR_=2.14×10^-19^) subfields, the INL-ELM outer (*d*=−0.42, *p*=0.00013, *p*_FDR_=0.00026), inner (*d*=−0.73, *p*=4.21×10^−11^, *p*_FDR_=2.40×10^−10^), and central (*d*=−0.86, *p*=7.11×10^−15^, *p*_FDR_=1.11×10^−13^) subfields, the ELM-ISOS outer (*d*=−0.47, *p*=2.05×10^−5^, *p*_FDR_=4.61×10^−5^), inner (*d*=−0.48, *p*=1.05×10^−5^, *p*_FDR_=2.45×10^−5^), and central (*d*=−0.64, *p*=1.99×10^−9^, *p*_FDR_=8.96×10^−9^) subfields, and the ISOS-RPE outer (*d*=−0.76, *p*=1.10×10^−11^, *p*_FDR_=8.67×10^−11^), inner (*d*=−0.65, *p*=6.91×10^−9^, *p*_FDR_=2.56×10^−8^), and central (*d*=−0.60, *p*=1.15×10^−7^, *p*_FDR_=3.44×10^−7^) subfields compared to HC.

However, participants with PSDs in cluster 1 did not differ from HCs except in the thickness of the RNFL, which was thicker in cluster 1 PSD (*d*=0.32, *p*=0.0059, *p*_FDR_=0.0093). The other layers did not differ in thickness (*p*>0.05).

***Differences in biopsychosocial variables***

Participants with PSD in cluster 2 differed significantly from those in cluster 1 on age (*d*=0.45, *p*=0.00063, *p*_FDR_=0.0043), race (*d*=0.50, *p*=9.63×10^−5^, *p*_FDR_=0.00077), household income (*d*=−1.12, *p*=2.22×10^−16^, *p*_FDR_=3.55×10^−15^), and TDI (*d*=0.71, *p*=1.61×10^−8^, *p*_FDR_=1.94×10^−7^), but did not differ with regard to sex (*d*=−0.0020, *p*=0.99, *p*_FDR_=0.99). Moreover, they did not differ with regard to health (BMI (*d*=0.39, *p*=0.28, *p*_FDR_=0.54), systolic blood pressure (*d*=0.34, *p*=0.35, *p*_FDR_=0.63), alcohol drinking (*d*=−0.28, *p*=0.44, *p*_FDR_=0.75), or tobacco smoking (*d*=0.18, *p*=0.10, *p*_FDR_=0.27)) or ocular metrics (visual acuity (*d*=0.54, *p*=0.14, *p*_FDR_=0.34), corneal hysteresis (*d*=−0.15, *p*=0.69, *p*_FDR_=0.97), ganglion cell intraocular-pressure (*d*=−0.28, *p*=0.44, *p*_FDR_=0.75), or corneal compensated intraocular-pressure (*d*=−0.0042, *p*=0.99, *p*_FDR_=0.99)). Although fluid intelligence was nominally significantly different between cluster 2 and cluster 1 (*d*=−0.75, *p*=0.045, *p*_FDR_=0.17), it did not survive FDR correction. Processing speed and reaction time (*d*=0.53, *p*=0.15, *p*_FDR_=0.34) and prospective memory (*d*=−0.70, *p*=0.059, *p*_FDR_=0.19) were also not different between cluster 2 and cluster 1.

Participants with PSD in cluster 2 also differed significantly from HCs with regard to race (*d*=0.27, *p*=0.0056, *p*_FDR_=0.034), household income (d=−1.35, pFDR=1.11×10−26), and TDI (d=0.93, pFDR= 2.41×10−15), but did not differ in age (*d*=0.19, *p*=0.070, *p*_FDR_=0.19) or sex (*d*=−0.00088, *p*=0.99, *p*_FDR_=0.99). They also differed in tobacco smoking (*d*=0.55, *p*=1.48×10^−6^, *p*_FDR_=1.42×10^−5^) and BMI (*d*=0.67, *p*=0.043, *p*_FDR_=0.17), but BMI did not survive FDR correction. Moreover, they did not differ with regard to systolic blood pressure (*d*=−0.027, *p*=0.93, *p*_FDR_=0.99) or alcohol drinking (*d*=−0.15, *p*=0.64, *p*_FDR_=0.97). They did not differ with regard to ocular measures (visual acuity (*d*=0.37, *p*=0.26, *p*_FDR_=0.52), corneal hysteresis (*d*=−0.12, *p*=0.72, *p*_FDR_=0.97), ganglion cell intraocular-pressure (*d*=0.024, *p*=0.94, *p*_FDR_=0.99), or corneal compensated intraocular-pressure (*d*=0.079, *p*=0.81, *p*_FDR_=0.97)). Similarly, cluster 2 PSD participants and HCs differed in fluid intelligence (*d*=−0.79, *p*=0.018, *p*_FDR_=0.088) but it did not survive FDR correction, while processing speed and reaction time (*d*=0.39, *p*=0.24, *p*_FDR_=0.50) and prospective memory (*d*=−0.63, *p*=0.058, *p*_FDR_=0.19) did not differ.

Participants with PSD in cluster 1 did not differ from HCs with regard to demographic (age, sex, race, household income, TDI; *p*>0.05), ocular (visual acuity, corneal hysteresis, ganglion cell intraocular-pressure, corneal compensated intraocular-pressure; *p*>0.05), and cognitive (processing speed and reaction time, fluid intelligence, and prospective memory; *p*>0.05) variables. While they also did not differ with regard to BMI (*d*=0.25, *p*=0.73, *p*_FDR_=0.77), systolic blood pressure (*d*=−0.36, *p*=0.29, *p*_FDR_=0.54), or alcohol drinking (*d*=0.14, *p*=0.68, *p*_FDR_=0.97), they did differ statistically with regard to tobacco smoking (*d*=0.36, *p*=0.0084, *p*_FDR_=0.045).

***Results of unimputed data***

*Canonical Correlation Analysis*

After removing participants who had any missing data from the variables used (retinal layers thickness, demographic, economic, health, ocular, and cognitive) and removing variables with a variance inflation factor>10 (ISOS-RPE inner thickness, INL-ELM inner thickness, and corneal compensated intraocular-pressure), we performed CCA. We scaled and centered the retinal layer thickness data and demographic, economic, health, ocular, and cognitive variables. Canonical correlation analysis produced 11 latent variate pairs, with the largest canonical correlation equaling 0.52. This first latent variate pair had a p-value < 0.0001 following permutation testing and was reliable following 10-fold cross-validation (mean training canonical correlation=0.517; mean testing canonical correlation=0.512). Latent variate pairs two (canonical correlation=0.42) and three (canonical correlation=0.31) were also significant via permutation testing (latent variate pair 2 *p*<0.0001; latent variate pair 3 *p*=0.0079) while the remaining 8 latent variate pairs were not (*p*>0.05). Latent variate pairs 2 and 3 showed very good reliability (latent variate pair 2 mean training canonical correlation=0.416; latent variate pair 2 mean testing canonical correlation=0.410; latent variate pair 3 mean training canonical correlation=0.313; latent variate pair 3 mean testing canonical correlation=0.309). While the remaining latent variate pairs also showed reliable canonical correlations via cross-validation, due to insignificant permutation tests (*p*>0.05) and smaller canonical correlations (*r*<0.26), we did not consider the remaining latent variate pairs for clustering.

*Hierarchical clustering*

For the first 3 latent variate pairs, Ward’s method produced the highest agglomerative coefficient (ac) (latent variate pair 1 Ward’s=0.994; latent variate pair 2 Ward’s=0.993; latent variate pair 1 Ward’s 3=0.993). While the other methods were also high (range=0.882—0.984), we used Ward’s method for clustering. Latent variate pair 1 obtained the highest silhouette score for two clusters (silhouette score=0.38), as did for latent variate pairs 2 (silhouette score=0.35) and 3 (silhouette score=0.31), though the silhouette scores were not as high. None of the 3 latent variate pairs produced significant p-values following significance testing (latent variate pair 1 *p*=0.61; latent variate pair 1 *p*=0.80; latent variate pair 1 *p*=0.62). None of the latent variate pairs clusters showed good stability following bootstrapping (latent variate pair 1 cluster 1 Jaccard index=0.73, latent variate pair 1 cluster 2 Jaccard index=0.66; latent variate pair 2 cluster 1 Jaccard index=0.60, latent variate pair 2 cluster 2 Jaccard index=0.48; latent variate pair 3 cluster 1 Jaccard index=0.64, latent variate pair 3 cluster 2 Jaccard index=0.70). In summary, while all 3 latent variate pairs showed moderate-sized, significant, and reliable canonical correlations, none produced significant or stable clusters.

Linkage methods across imputations

Ward’s linkage method produced the highest ac for clustering in all 10 imputations (mean ac=0.996; median ac=0.996; minimum ac=0.995; maximum ac=0.996), compared to the average (mean ac=0.969; median ac=0.970; minimum ac=0.958; maximum ac=0.974), single (mean ac=0.939; median ac=0.944; minimum ac=0.900; maximum ac=0.954), and complete (mean ac=0.984; median ac=0.984; minimum ac=0.983; maximum ac=0.985) linkage methods. Note that the maximum ac for the average, single, and complete linkage methods are not greater than the minimum ac for Ward’s method. Therefore, Ward’s linkage method was used for clustering in all 10 imputed datasets.

Retinal layer thickness cluster differences covaried models

When covarying for age and comparing participants with PSDs in cluster 2 to cluster 1, those in cluster 2 had significantly thinner macular (*d*=−0.77, *p*=1.15×10^−8^, *p*_FDR_=4.25×10^−8^), RNFL (*d*=−0.66, *p*=2.70×10^−7^, *p*_FDR_=8.10×10^−7^), INL-RPE (*d*=−0.81, *p*=9.02×10^−10^, *p*_FDR_=4.37×10^−9^), INL-ELM (*d*=−0.51, *p*=8.63×10^−5^, *p*_FDR_=0.00018), ELM-ISOS (*d*=−0.62, *p*=5.35×10^−6^, *p*_FDR_=1.47×10^−5^), and ISOS-RPE (*d*=−0.52, *p*= 5.98×10^−5^, *p*_FDR_=0.00013) layers and significantly thicker RPE layer (*d*=0.36, *p*=0.0033, *p*_FDR_=0.0058). When examining subfields of photoreceptor layers, cluster 2 also had thinner INL-RPE outer (*d*=−0.73, *p*=3.21×10^−8^, *p*_FDR_=1.12×10^−7^), inner (*d*=−0.87, *p*=4.37×10^−11^, *p*_FDR_=2.50×10^−10^), and central (*d*=−0.93, *p*=1.91×10^−12^, *p*_FDR_=1.72×10^−11^) subfields, thinner INL-ELM outer (*d*=−0.39, *p*=0.0026, *p*_FDR_=0.0047), inner (*d*=−0.71, *p*=4.18×10^−8^, *p*_FDR_=1.39×10^−7^), and central (*d*=−0.79, *p*=1.61×10^−9^, *p*_FDR_=1.09×10^−8^) subfields, thinner ELM-ISOS outer (*d*=−0.57, *p*=3.13×10^−5^, *p*_FDR_=7.31×10^−5^), inner (*d*=−0.56, *p*=3.84×10^−5^, *p*_FDR_=8.65×10^−5^), and central (*d*=−0.83, *p*=8.35×10^−10^, *p*_FDR_=4.37×10^−9^) subfields, and thinner ISOS-RPE outer (*d*=−0.54, *p*=2.76×10^−5^, *p*_FDR_=6.46×10^−5^), inner (*d*=−0.41, *p*=0.0016, *p*_FDR_=0.0029), and central (*d*=−0.38, *p*=0.0043, *p*_FDR_=0.0071) subfields layer thicknesses compared to cluster 1. The GC-IPL (*d*=−0.12, *p*=0.35, *p*_FDR_=0.44) and INL (*d*=−0.12, *p*=0.37, *p*_FDR_=0.44) layers were not different between cluster 2 and cluster 1.

When covarying for race and comparing participants with PSDs in cluster 2 to cluster 1, those in cluster 2 had significantly thinner macular (*d*=−0.74, *p*=4.05×10^−8^, *p*_FDR_=1.59×10^−7^), RNFL (*d*=−0.66, *p*=2.40×10^−7^, *p*_FDR_=7.87×10^−7^), INL-RPE (*d*=−0.75, *p*=1.02×10^−8^, *p*_FDR_=4.28×10^−8^), INL-ELM (*d*=−0.43, *p*=0.00086, *p*_FDR_=0.0016), ELM-ISOS (*d*=−0.69, *p*=3.96×10^−7^, *p*_FDR_=1.19×10^−6^), and ISOS-RPE (*d*=−0.50, *p*= 8.75×10^−5^, *p*_FDR_=0.00018) layers and significantly thicker RPE layer (*d*=0.31, *p*=0.0097, *p*_FDR_=0.016). When examining subfields of photoreceptor layers, cluster 2 also had thinner INL-RPE outer (*d*=−0.67, *p*=2.50×10^−7^, *p*_FDR_=7.87×10^−7^), inner (*d*=−0.81, *p*=6.21×10^−10^, *p*_FDR_=3.56×10^−9^), and central (*d*=−0.86, *p*=4.77×10^−11^, *p*_FDR_=3.34×10^−10^) subfields, thinner INL-ELM outer (*d*=−0.32, *p*=0.013, *p*_FDR_=0.021), inner (*d*=−0.62, *p*=1.30×10^−6^, *p*_FDR_=3.72×10^−6^), and central (*d*=−0.70, *p*=9.43×10^−8^, *p*_FDR_=3.49×10^−7^) subfields, thinner ELM-ISOS outer (*d*=−0.64, *p*=2.29×10^−6^, *p*_FDR_=6.02×10^−6^), inner (*d*=−0.61, *p*=9.41×10^−6^, *p*_FDR_=2.28×10^−5^), and central (*d*=−0.80, *p*=1.91×10^−9^, *p*_FDR_=1.00×10^−8^) subfields, and thinner ISOS-RPE outer (*d*=−0.52, *p*=5.44×10^−5^, *p*_FDR_=0.00012), inner (*d*=−0.41, *p*=0.0012, *p*_FDR_=0.0022), and central (*d*=−0.40, *p*=0.0024, *p*_FDR_=0.0043) subfields layer thicknesses compared to cluster 1. The GC-IPL (*d*=−0.12, *p*=0.35, *p*_FDR_=0.40) and INL (*d*=−0.10, *p*=0.41, *p*_FDR_=0.46) layers were not different between cluster 2 and cluster 1.

Then, when covarying for age and race and comparing participants with PSDs in cluster 2 to cluster 1, those in cluster 2 had significantly thinner macular (*d*=−0.68, *p*=2.47×10^−7^, *p*_FDR_=8.65×10^−7^), RNFL (*d*=−0.62, *p*=6.48×10^−7^, *p*_FDR_=2.04×10^−6^), INL-RPE (*d*=−0.72, *p*=1.74×10^−8^, *p*_FDR_=7.85×10^−8^), INL-ELM (*d*=−0.42, *p*=0.00076, *p*_FDR_=0.0015), ELM-ISOS (*d*=−0.62, *p*=3.55×10^−6^, *p*_FDR_=1.02×10^−5^), and ISOS-RPE (*d*=−0.49, *p*=0.00011, *p*_FDR_=0.00022) layers and significantly thicker RPE layer (*d*=0.32, *p*=0.0071, *p*_FDR_=0.012). When examining subfields of photoreceptor layers, cluster 2 also had thinner INL-RPE outer (*d*=−0.65, *p*=4.04×10^−7^, *p*_FDR_=1.34×10^−6^), inner (*d*=−0.78, *p*=1.02×10^−9^, *p*_FDR_=5.82×10^−9^), and central (*d*=−0.83, *p*=9.71×10^−11^, *p*_FDR_=6.79×10^−10^) subfields, thinner INL-ELM outer (*d*=−0.31, *p*=0.013, *p*_FDR_=0.021), inner (*d*=−0.63, *p*=8.54×10^−7^, *p*_FDR_=2.56×10^−6^), and central (*d*=−0.69, *p*=7.19×10^−8^, *p*_FDR_=2.83×10^−7^) subfields, thinner ELM-ISOS outer (*d*=−0.58, *p*=1.50×10^−5^, *p*_FDR_=3.78×10^−5^), inner (*d*=−0.53, *p*=7.68×10^−5^, *p*_FDR_=0.00017), and central (*d*=−0.75, *p*=1.07×10^−8^, *p*_FDR_=5.64×10^−8^) subfields, and thinner ISOS-RPE outer (*d*=−0.51, *p*=5.53×10^−5^, *p*_FDR_=0.00012), inner (*d*=−0.39, *p*=0.0018, *p*_FDR_=0.0034), and central (*d*=−0.36, *p*=0.0045, *p*_FDR_=0.0081) subfields layer thicknesses compared to cluster 1. The GC-IPL (*d*=−0.06, *p*=0.63, *p*_FDR_=0.67) and INL (*d*=−0.08, *p*=0.51, *p*_FDR_=0.58) layers were not different between cluster 2 and cluster 1.

When covarying for age, participants with PSDs in cluster 2 had significantly thinner macular (*d*=−0.78, *p*=1.31×10^−12^, *p*_FDR_=1.38×10^−11^), RNFL (*d*=−0.37, *p*=0.00062, *p*_FDR_=0.0012), GC-IPL (*d*=−0.28, *p*=0.0084, *p*_FDR_=0.013), INL-RPE (*d*=−0.92, *p*=2.22×10^−16^, *p*_FDR_=4.66×10^−15^), INL-ELM, (*d*=−0.54, *p*=7.67×10^−7^, *p*_FDR_=2.20×10^−6^), and ELM-ISOS (*d*=−0.49, *p*=7.03×10^−6^, *p*_FDR_=1.84×10^−5^), ISOS-RPE (*d*=−0.74, *p*=2.39×10^−11^, *p*_FDR_=1.64×10^−10^) and thicker RPE layers (*d*=0.31, *p*=0.0039, *p*_FDR_=0.0067) compared to HCs, while the INL layer was not different (*d*=−0.17, *p*=0.12, *p*_FDR_=0.17). When examining subfields of photoreceptor layers, cluster 2 also had thinner layer thicknesses for the INL-RPE outer (*d*=−0.83, *p*=9.95×10^−14^, *p*_FDR_=1.25×10^−12^), inner (*d*=−0.98, *p*=1.86×10^−18^, *p*_FDR_=5.86×10^−17^), and central (*d*=−1.05, *p*=4.33×10^−21^, *p*_FDR_=2.73×10^−19^) subfields, the INL-ELM outer (*d*=−0.42, *p*=0.00012, *p*_FDR_=0.00023), inner (*d*=−0.74, *p*=2.60×10^−11^, *p*_FDR_=1.64×10^−10^), and central (*d*=−0.86, *p*=4.66×10^−15^, *p*_FDR_=7.34×10^−14^) subfields, the ELM-ISOS outer (*d*=−0.45, *p*=4.48×10^−5^, *p*_FDR_=9.73×10^−5^), inner (*d*=−0.45, *p*=2.61×10^−5^, *p*_FDR_=6.46×10^−5^), and central (*d*=−0.63, *p*=3.83×10^−9^, *p*_FDR_=1.61×10^−8^) subfields, and the ISOS-RPE outer (*d*=−0.75, *p*=1.09×10^−11^, *p*_FDR_=8.56×10^−11^), inner (*d*=−0.64, *p*=9.76×10^−9^, *p*_FDR_=3.84×10^−8^), and central (*d*=−0.58, *p*=1.91×10^−7^, *p*_FDR_=6.03×10^−7^) subfields compared to HCs.

Next, when covarying for race, participants with PSDs in cluster 2 had significantly thinner macular (*d*=−0.76, *p*=5.09×10^−12^, *p*_FDR_=5.35×10^−11^), RNFL (*d*=−0.37, *p*=0.00063, *p*_FDR_=0.0012), GC-IPL (*d*=−0.27, *p*=0.011, *p*_FDR_=0.0018), INL-RPE (*d*=−0.88, *p*=2.00×10^−15^, *p*_FDR_=4.20×10^−14^), INL-ELM, (*d*=−0.50, *p*=5.30×10^−6^, *p*_FDR_=1.34×10^−5^), and ELM-ISOS (*d*=−0.52, *p*=1.72×10^−6^, *p*_FDR_=4.70×10^−6^), ISOS-RPE (*d*=−0.73, *p*=4.05×10^−11^, *p*_FDR_=3.19×10^−10^) and thicker RPE layers (*d*=0.28, *p*=0.0080, *p*_FDR_=0.014) compared to HCs, while the INL layer was not different (*d*=−0.16, *p*=0.14, *p*_FDR_=0.20). When examining subfields of photoreceptor layers, cluster 2 also had thinner layer thicknesses for the INL-RPE outer (*d*=−0.80, *p*=6.61×10^−13^, *p*_FDR_=8.33×10^−12^), inner (*d*=−0.95, *p*=1.95×10^−17^, *p*_FDR_=6.14×10^−16^), and central (*d*=−1.01, *p*=6.82×10^−20^, *p*_FDR_=4.30×10^−18^) subfields, the INL-ELM outer (*d*=−0.38, *p*=0.00048, *p*_FDR_=0.00098), inner (*d*=−0.68, *p*=3.98×10^−10^, *p*_FDR_=2.51×10^−9^), and central (*d*=−0.81, *p*=1.30×10^−13^, *p*_FDR_=2.05×10^−12^) subfields, the ELM-ISOS outer (*d*=−0.48, *p*=1.03×10^−5^, *p*_FDR_=2.39×10^−5^), inner (*d*=−0.46, *p*=1.43×10^−5^, *p*_FDR_=3.21×10^−5^), and central (*d*=−0.61, *p*=8.50×10^−9^, *p*_FDR_=3.88×10^−8^) subfields, and the ISOS-RPE outer (*d*=−0.74, *p*=2.50×10^−11^, *p*_FDR_=2.25×10^−10^), inner (*d*=−0.64, *p*=8.62×10^−9^, *p*_FDR_=3.88×10^−8^), and central (*d*=−0.59, *p*=1.22×10^−7^, *p*_FDR_=4.28×10^−7^) subfields compared to HCs.

When covarying for age and race, participants with PSDs in cluster 2 had significantly thinner macular (*d*=−0.73, *p*=1.54×10^−11^, *p*_FDR_=1.62×10^−10^), RNFL (*d*=−0.35, *p*=0.00084, *p*_FDR_=0.0016), GC-IPL (*d*=−0.25, *p*=0.021, *p*_FDR_=0.033), INL-RPE (*d*=−0.87, *p*=2.89×10^−15^, *p*_FDR_=6.06×10^−14^), INL-ELM, (*d*=−0.50, *p*=4.79×10^−6^, *p*_FDR_=1.28×10^−5^), and ELM-ISOS (*d*=−0.49, *p*=4.86×10^−6^, *p*_FDR_=1.28×10^−5^), ISOS-RPE (*d*=−0.72, *p*=4.84×10^−11^, *p*_FDR_=3.81×10^−10^) and thicker RPE layers (*d*=0.29, *p*=0.0065, *p*_FDR_=0.011) compared to HCs, while the INL layer was not different (*d*=−0.15, *p*=0.17, *p*_FDR_=0.23). When examining subfields of photoreceptor layers, cluster 2 also had thinner layer thicknesses for the INL-RPE outer (*d*=−0.78, *p*=9.15×10^−13^, *p*_FDR_=1.15×10^−11^), inner (*d*=−0.93, *p*=2.66×10^−17^, *p*_FDR_=8.39×10^−16^), and central (*d*=−0.99, *p*=1.094×10^−19^, *p*_FDR_=6.89×10^−18^) subfields, the INL-ELM outer (*d*=−0.38, *p*=0.00047, *p*_FDR_=0.00095), inner (*d*=−0.69, *p*=2.81×10^−10^, *p*_FDR_=1.77×10^−9^), and central (*d*=−0.81, *p*=1.01×10^−13^, *p*_FDR_=1.59×10^−12^) subfields, the ELM-ISOS outer (*d*=−0.45, *p*=2.50×10^−5^, *p*_FDR_=6.05×10^−5^), inner (*d*=−0.43, *p*=4.07×10^−5^, *p*_FDR_=9.49×10^−5^), and central (*d*=−0.59, *p*=1.88×10^−8^, *p*_FDR_=7.91×10^−8^) subfields, and the ISOS-RPE outer (*d*=−0.73, *p*=2.57×10^−11^, *p*_FDR_=2.31×10^−10^), inner (*d*=−0.63, *p*=1.28×10^−8^, *p*_FDR_=6.20×10^−8^), and central (*d*=−0.58, *p*=2.19×10^−7^, *p*_FDR_=8.10×10^−7^) subfields compared to HCs.

However, when covarying for age, race, and age and race, participants with PSDs in cluster 1 again did not differ from HCs except for the RNFL layer thickness. It was thicker in cluster 1 PSD individuals when adjusting for age (*d*=0.31, *p*=0.0075, *p*_FDR_=0.012), race (*d*=0.31, *p*=0.0067), and age and race (*d*=0.29, *p*=0.0087). The other layers did not differ in thickness (*p*>0.05).

Biopsychosocial comparisons results when covarying for age, race, and age and race.

When covarying just for age, participants with PSDs in cluster 2 differed significantly from those in cluster 1 race (*d*=0.53, *p*=3.18×10^−5^, *p*_FDR_=0.00027), household income (*d*=−1.06, *p*=2.22×10^−15^, *p*_FDR_=3.77×10^−14^), and TDI (*d*=0.79, *p*=2.43×10^−10^, *p*_FDR_=3.09×10^−9^), but did not differ with regard to sex (*d*=0.000048, *p*=0.99, *p*_FDR_=0.99). Moreover, they did not differ with regard to health (BMI (*d*=0.41, *p*=0.26, *p*_FDR_=0.56), systolic blood pressure (*d*=0.23, *p*=0.53, *p*_FDR_=0.84), alcohol drinking (*d*=−0.26, *p*=0.47, *p*_FDR_=0.75), or tobacco smoking (*d*=0.23, *p*=0.035, *p*_FDR_=0.15)) or ocular metrics (visual acuity (*d*=0.46, *p*=0.20, *p*_FDR_=0.47), corneal hysteresis (*d*=−0.12, *p*=0.73, *p*_FDR_=0.94), ganglion cell intraocular-pressure (*d*=−0.15, *p*=0.68, *p*_FDR_=0.94), or corneal compensated intraocular-pressure (*d*=−0.064, *p*=0.86, *p*_FDR_=0.99)). Again, fluid intelligence was nominally significantly different between cluster 2 and cluster 1 (*d*=−0.74, *p*=0.044, *p*_FDR_=0.15) but did not survive FDR correction. Processing speed and reaction time (*d*=0.39, *p*=0.28, *p*_FDR_=0.58) and prospective memory (*d*=−0.66, *p*=0.072, *p*_FDR_=0.19) were also not different between cluster 2 and cluster 1.

When covarying just for race, participants with PSDs in cluster 2 differed significantly from those in cluster 1 age (*d*=0.49, *p*=0.00021, *p*_FDR_=0.0046), household income (*d*=−1.12, *p*=3.38×10^−17^, *p*_FDR_=3.77×10^−14^), and TDI (*d*=0.62, *p*=4.00×10^−7^, *p*_FDR_=3.09×10^−9^), but did not differ with regard to sex (*d*=0.010, *p*=0.94, *p*_FDR_=0.99). They did not differ with regard to health (BMI (*d*=0.38, *p*=0.29, *p*_FDR_=0.56), systolic blood pressure (*d*=0.33, *p*=0.36, *p*_FDR_=0.84), alcohol drinking (*d*=−0.27, *p*=0.46, *p*_FDR_=0.79), or tobacco smoking (*d*=0.15, *p*=0.15, *p*_FDR_=0.14)), ocular metrics (visual acuity (*d*=0.58, *p*=0.11, *p*_FDR_=0.47), corneal hysteresis (*d*=−0.058, *p*=0.87, *p*_FDR_=0.94), ganglion cell intraocular-pressure (*d*=−0.047, *p*=0.90, *p*_FDR_=0.94), or corneal compensated intraocular-pressure (*d*=−0.017, *p*=0.96, *p*_FDR_=0.99)), or cognitive measures (Fluid intelligence (*d*=−0.59, *p*=0.11, *p*_FDR_=0.15), processing speed and reaction time (*d*=0.44, *p*=0.23, *p*_FDR_=0.58) and prospective memory (*d*=−0.57, *p*=0.12, *p*_FDR_=0.19)).

Next, when covarying for age and race, participants with PSDs in cluster 2 differed significantly from those in cluster 1 regarding household income (*d*=−1.06, *p*=4.44×10^−16^, *p*_FDR_=6.66×10^−15^), and TDI (*d*=0.70, *p*=8.87×10^−9^, *p*_FDR_=9.98×10^−8^), but did not differ with regard to sex (*d*=0.013, *p*=0.92, *p*_FDR_=0.98). They did not differ with regard to health (BMI (*d*=0.40, *p*=0.27, *p*_FDR_=0.65), systolic blood pressure (*d*=0.21, *p*=0.56, *p*_FDR_=0.97), alcohol drinking (*d*=−0.24, *p*=0.51, *p*_FDR_=0.96), or tobacco smoking (*d*=0.20, *p*=0.054, *p*_FDR_=0.27)), ocular metrics (visual acuity (*d*=0.50, *p*=0.17, *p*_FDR_=0.48), corneal hysteresis (*d*=−0.022, *p*=0.95, *p*_FDR_=0.98), ganglion cell intraocular-pressure (*d*=−0.11, *p*=0.76, *p*_FDR_=0.98), or corneal compensated intraocular-pressure (*d*=−0.087, *p*=0.81, *p*_FDR_=0.98)), or cognitive measures (Fluid intelligence (*d*=−0.56, *p*=0.13, *p*_FDR_=0.48), processing speed and reaction time (*d*=0.27, *p*=0.46, *p*_FDR_=0.89) and prospective memory (*d*=−0.52, *p*=0.16, *p*_FDR_=0.48)).

When covarying for age, participants with PSD in cluster 2 also differed significantly from HCs with regard to race (*d*=0.28, *p*=0.0034, *p*_FDR_=0.022), household income (*d*=−1.33, *p*=8.19×10^−28^, *p*_FDR_=4.18×10^−26^) and TDI (*d*=0.98, *p*=2.16×10^−18^, *p*_FDR_=5.5×10^−17^), but did not differ in sex (*d*=0.000021, *p*=0.99, *p*_FDR_=0.99). They also differed in tobacco smoking (*d*=0.58, *p*=3.71×10^−7^, *p*_FDR_=3.78×10^−6^) and BMI (*d*=0.68, *p*=0.041, *p*_FDR_=0.15), but BMI did not survive FDR correction. Moreover, they did not differ with regard to systolic blood pressure (*d*=−0.078, *p*=0.81, *p*_FDR_=0.99) or alcohol drinking (*d*=−0.15, *p*=0.66, *p*_FDR_=0.94). They did not differ with regard to ocular measures (visual acuity (*d*=0.34, *p*=0.30, *p*_FDR_=0.58), corneal hysteresis (*d*=−0.11, *p*=0.74, *p*_FDR_=0.94), ganglion cell intraocular-pressure (*d*=−0.0012, *p*=0.99, *p*_FDR_=0.99), or corneal compensated intraocular-pressure (*d*=0.053, *p*=0.87, *p*_FDR_=0.99)). Similarly, cluster 2 PSD participants and HCs differed in fluid intelligence (*d*=−0.79, *p*=0.019, *p*_FDR_=0.096) but it did not survive FDR correction, while processing speed and reaction time (*d*=0.34, *p*=0.31, *p*_FDR_=0.58) and prospective memory (*d*=−0.62, *p*=0.064, *p*_FDR_=0.19) did not differ.

When covarying for race, participants with PSD in cluster 2 also differed significantly from HCs with regard to household income (*d*=−1.35, *p*=2.86×10^−29^, *p*_FDR_=4.18×10^−26^) and TDI (*d*=0.88, *p*=1.78×10^−15^, *p*_FDR_=5.5×10^−17^) but did not differ in sex (*d*=0.0064, *p*=0.96, *p*_FDR_=0.99). Age was nominally different between cluster 2 PSD participants and HCs, but this did not survive FDR correction (*d*=0.21, *p*=0.040, *p*_FDR_=0.19). They also differed in tobacco smoking (*d*=0.53, *p*=3.33×10^−6^, *p*_FDR_=3.78×10^−6^) but did not differ regarding BMI (*d*=0.64, *p*=0.054, *p*_FDR_=0.15), systolic blood pressure (*d*=−0.039, *p*=0.91, *p*_FDR_=0.99) or alcohol drinking (*d*=−0.16, *p*=0.63, *p*_FDR_=0.94). They did not differ with regard to ocular measures (visual acuity (*d*=0.40, *p*=0.22, *p*_FDR_=0.58), corneal hysteresis (*d*=−0.077, *p*=0.82, *p*_FDR_=0.94), ganglion cell intraocular-pressure (*d*=−0.051, *p*=0.88, *p*_FDR_=0.99), or corneal compensated intraocular-pressure (*d*=0.069, *p*=0.83, *p*_FDR_=0.99)). Similarly, cluster 2 PSD participants and HCs differed in fluid intelligence (*d*=−0.70, *p*=0.038, *p*_FDR_=0.096) but it did not survive FDR correction, while processing speed and reaction time (*d*=0.33, *p*=0.32, *p*_FDR_=0.58) and prospective memory (*d*=−0.55, *p*=0.10, *p*_FDR_=0.19) did not differ.

When covarying for age and race, participants with PSD in cluster 2 also differed significantly from HCs with regard to household income (*d*=−1.33, *p*=1.58×10^−28^, *p*_FDR_=7.13×10^−27^) and TDI (*d*=0.93, *p*=4.74×10^−17^, *p*_FDR_=1.07×10^−15^) but did not differ in sex (*d*=0.0076, *p*=0.94, *p*_FDR_=0.98). They also differed in tobacco smoking (*d*=0.56, *p*=7.92×10^−7^, *p*_FDR_=7.13×10^−6^) but did not differ regarding BMI (*d*=0.65, *p*=0.0501, *p*_FDR_=0.27), systolic blood pressure (*d*=−0.10, *p*=0.75, *p*_FDR_=0.98) or alcohol drinking (*d*=−0.14, *p*=0.17, *p*_FDR_=0.48). They did not differ with regard to ocular measures (visual acuity (*d*=0.36, *p*=0.27, *p*_FDR_=0.65), corneal hysteresis (*d*=−0.065, *p*=0.84, *p*_FDR_=0.98), ganglion cell intraocular-pressure (*d*=0.023, *p*=0.95, *p*_FDR_=0.98), or corneal compensated intraocular-pressure (*d*=0.037, *p*=0.91, *p*_FDR_=0.98)). Similarly, cluster 2 PSD participants and HCs differed in fluid intelligence (*d*=−0.68, *p*=0.041, *p*_FDR_=0.27) but it did not survive FDR correction, while processing speed and reaction time (*d*=0.26, *p*=0.44, *p*_FDR_=0.89) and prospective memory (*d*=−0.52, *p*=0.11, *p*_FDR_=0.47) did not differ.

And when adjusting for age, participants with PSD in cluster 1 differed from HCs with regard to race (*d*=−0.27, *p*=0.045, *p*_FDR_=0.15) but did not survive FDR correction. Other demographic measures (sex, household income, TDI) did not differ (*p*>0.05). They also differed with regard to tobacco smoking (*d*=0.32, *p*=0.019, *p*_FDR_=0.10) but again, this comparison did not survive FDR correction, and the others were not significant either (BMI, systolic blood pressure, alcohol drinking; *p*>0.05). In the same way, the ocular (visual acuity, corneal hysteresis, ganglion cell intraocular-pressure, corneal compensated intraocular-pressure) and cognitive measures (processing speed and reaction time, fluid intelligence, and prospective memory) did not differ (*p*>0.05). When covarying for race, age (*d*=−0.27, *p*=0.021, *p*_FDR_=0.15) and tobacco smoking (*d*=0.37, *p*=0.0067, *p*_FDR_=0.10) were also nominally significant but did not survive FDR correction, while other comparisons between participants with PSDs in cluster 1 and HCs were not significantly different (*p*>0.05). Lastly, when covarying for both age and race, only tobacco smoking was nominally significant (*d*=0.34, *p*=0.016, *p*_FDR_=0.12) and this correction did not survive FDR correction. No other comparisons were significant (*p*>0.05).
